# Biomathematical models for genetic diversity analyses in complete genomes of SARS-CoV-2

**DOI:** 10.1101/2020.10.01.20205120

**Authors:** Robson da Silva Ramos, Pierre Teodósio Felix, Dallynne Bárbara Ramos Venâncio, Cícero Batista do Nascimento Filho, Antônio João Paulino

## Abstract

In this work, we evaluated the levels of genetic diversity in 38 complete genomes of SARS-CoV-2, publicly available on the National Center for Biotechnology Information (NCBI) platform and from six countries in South America (Brazil, Chile, Peru, Colombia, Uruguay and Venezuela with 16, 11, 1, 1, 1, 7 haplotypes, respectively), all with an extension of 29,906 bp and Phred values ≥ 40. These haplotypes were previously used for phylogenetic analyses, following the alignment protocols of the MEGA X software; where all gaps and unconserved sites were extracted for the construction of phylogenetic trees. The specific methodologies for Paired F_ST_ estimators, Molecular Variance (AMOVA), Genetic Distance, mismatch, demographic and spatial expansion analyses, molecular diversity and evolutionary divergence time analyses, were obtained using 20,000 random permutation.

## 1. Methodology

**Databank:** The 38 complete genome sequences of SARS-CoV-2 from South America (Brazil, Chile, Peru, Colombia, Uruguay and Venezuela with 16, 11, 1, 2, 1, 7 haplotypes, respectively) all with 29,906 pb extension and Phred values ≥ 40 and which now make up our study PopSet, were recovered from GENBANK (https://www.ncbi.nlm.nih.gov/labs/virus/vssi/#/virus?SeqType_s=Nucleotide&VirusLineage_ss=SARS-CoV-2,%20taxid:2697049&Completeness_s=complete&Region_s=South%20America) on August 21, 2020).

**Phylogenetic analyses:** Nucleotide sequences previously described were used for phylogenetic analyses. The sequences were aligned using the MEGA X program (TAMURA *et al*., 2018) and the gaps were extracted for the construction of phylogenetic trees.

**Genetic Structuring Analyses**: Paired F_ST_ estimators, Molecular Variance (AMOVA), Genetic Distance, mismatch, demographic and spatial expansion analyses, molecular diversity and evolutionary divergence time were obtained with the Software Arlequin v. 3.5 (EXCOFFIER *et al*., 2005) using 1000 random permutations (NEI and KUMAR, 2000). The F_ST_ and geographic distance matrices were not compared. All steps of this process are described below.

### FOR GENETIC DIVERSITY

Among the routines of LaBECom, this test is used to measure the genetic diversity that is equivalent to the heterozygosity expected in the groups studied. We used for this the standard index of genetic diversity H, described by Nei (1987). Which can also be estimated by the method proposed by PONS and PETIT (1995).

### FOR SITE FREQUENCY SPECTRUM (SFS)

According to LaBECom protocols, we used this local frequency spectrum analytical test (SFS), from DNA sequence data that allows us to estimate the demographic parameters of the frequency spectrum. Simulations are made using fastsimcoal2 software, available in http://cmpg.unibe.ch/software/fastsimcoal2/.

### FOR MOLECULAR DIVERSITY INDICES

Molecular diversity indices are obtained by means of the average number of paired differences, as described by Tajima in 1993, in this test we used sequences that do not fit the model of neutral theory that establishes the existence of a balance between mutation and genetic drift.

### FOR CALCULATING THETA ESTIMATORs

Theta population parameters are used in our Laboratory when we want to qualify the genetic diversity of the populations studied. These estimates, classified as Theta Hom – which aim to estimate the expected homozygosity in a population in equilibrium between drift and mutation and the estimates Theta (S) (WATTERSON, 1975), Theta (K) (EWENS, 1972) and Theta (π) (TAJIMA, 1983).

### FOR THE CALCULATION OF The DISTRIBUTION OF MISMATCH

In LaBECom, analyses of the mismatch distribution are always performed relating the observed number of differences between haplotype pairs, trying to define or establish a pattern of population demographic behavior, as already described by (ROGERS; HARPENDING, 1992; Hudson, Hudson, HUDSON, SLATKIN, 1991; RAY et al., 2003, EXCOFFIER, 2004).

### FOR PURE DEMOGRAPHIC EXPANSION

This model is always used when we intend to estimate the probability of differences observed between two haplotypes not recombined and randomly chosen, this methodology in our laboratory is used when we assume that the expansion, in a haploid population, reached a momentary balance even having passed through τ generations, of sizes 0 N to 1 N. In this case, the probability of observing the S differences between two non-recombined and randomly chosen haplotypes is given by the probability of observing two haplotypes with S differences in this population (Watterson, 1975).

### FOR SPATIAL EXPANSION

The use of this model in LaBECom is usually indicated if the reach of a population is initially restricted to a very small area, and when one notices signs of a growth of the same, in the same space and over a relatively short time. The resulting population generally becomes subdivided in the sense that individuals tend to mate with geographically close individuals rather than random individuals. To follow the dimensions of spatial expansion, we at LaBECom always take into account:

L: Number of loci

Gamma Correction: This fix is always used when mutation rates do not seem uniform for all sites.

nd: Number of substitutions observed between two DNA sequences.

ns: Number of transitions observed between two DNA sequences.

nv: Number of transversions observed between two DNA sequences.

ω: G + C ratio, calculated in all DNA sequences of a given sample.

Paired Difference: Shows the number of loci for which two haplotypes are different.

Percentage difference: This difference is responsible for producing the percentage of loci for which two haplotypes are different.

### FOR HAPLOTYPIC INFERENCES

We use these inferences for haplotypic or genotypic data with unknown gametic phase. Following our protocol, inferences are estimated by observing the relationship between haplotype i and xi times its number of copies, generating an estimated frequency (^pi). With genotypic data with unknown gametic phase, the frequencies of haplotypes are estimated by the maximum likelihood method, and can also be estimated using the expected Maximization (MS) algorithm.

### FOR THE METHOD OF JUKES AND CANTOR

This method, when used in LaBECom, allows estimating a corrected percentage of how different two haplotypes are. This correction allows us to assume that there have been several substitutions per site, since the most recent ancestor of the two haplotypes studied. Here, we also assume a correction for identical replacement rates for all four nucleotides A C, G and T.

### FOR KIMURA METHOD WITH TWO PARAMETERS

Much like the previous test, this fix allows for multiple site substitutions, but takes into account different replacement rates between transitions and transversions.

### FOR TAMURA METHOD

We at LaBECom understand this method as an extension of the 2-parameter Kimura method, which also allows the estimation of frequencies for different haplotypes. However, transition-transversion relationships as well as general nucleotide frequencies are calculated from the original data.

### FOR The TAJIMA AND NEI METHOD

At this stage, we were also able to produce a corrected percentage of nucleotides for which two haplotypes are different, but this correction is an extension of the Jukes and Cantor method, with the difference of being able to do this from the original data.

### FOR TAMURA AND NEI MODEL

As in kimura’s models 2 parameters a distance of Tajima and Nei, this correction allows, inferring different rates of transversions and transitions, besides being able to distinguish transition rates between purines and pyrimidines.

### FOR ESTIMATING DISTANCES BETWEEN HAPLOTYPES PRODUCED BY RFLP

We use this method in our laboratory when we need to verify the number of paired differences scouting the number of different alleles between two haplotypes generated by RFLP.

### TO ESTIMATE DISTANCES BETWEEN HAPLOTYPES PRODUCED MICROSATELLITES

In this case, what applies is a simple count of the number of different alleles between two haplotypes. Using the sum of the square of the differences of repeated sites between two haplotypes (Slatkin, 1995).

### MINIMUM SPANNING NETWORK

To calculate the distance between OTU (operational taxonomic units) from the paired distance matrix of haplotypes, we used a Minimum Spanning Network (MSN) tree, with a slight modification of the algorithm described in Rohlf (1973). We usually use free software written in Pascal called MINSPNET. EXE running in DOS language, previously available at: http://anthropologie.unige.ch/LGB/software/win/min-span-net/.

### FOR GENOTYPIC DATA WITH UNKNOWN GAMETIC PHASE

#### EM algorithm

To estimate haplotypic frequencies we used the maximum likelihood model with an algorithm that maximizes the expected values. The use of this algorithm in LaBECom, allows to obtain the maximum likelihood estimates from multilocal data of gamtic phase is unknown (phenotypic data). It is a slightly more complex procedure since it does not allow us to do a simple gene count, since individuals in a population can be heterozygous to more than one locus.

#### ELB algorithm

Very similar to the previous algorithm, ELB attempts to reconstruct the gametic phase (unknown) of multilocal genotypes by adjusting the sizes and locations of neighboring loci to explore some rare recombination.

### FOR NEUTRALITY TESTS

#### Ewens-Watterson homozygosis test

We use this test in LaBECom for both haploid and diploid data. This test is used only as a way to summarize the distribution of allelic frequency, without taking into account its biological significance. This test is based on the sampling theory of neutral alllinks from Ewens (1972) and tested by Watterson (1978). It is now limited to sample sizes of 2,000 genes or less and 1,000 different alleles (haplotypes) or less. It is still used to test the hypothesis of selective neutrality and population balance against natural selection or the presence of some advantageous alleles.

#### Accurate Ewens-Watterson-Slatkin Test

This test created by Slatikin in 1994 and adapted by himself in 1996. is used in our protocols when we want to compare the probabilities of random samples with those of observed samples.

#### Chakraborty’s test of population amalgamation

This test was proposed by Chakrabordy in 1990, serves to calculate the observed probability of a randomly neutral sample with a number of alleles equal to or greater than that observed, it is based on the infinite allele model and sampling theory for neutral Alleles of Ewens (1972).

#### Tajima Selective Neutrality Test

We use this test in our Laboratory when DNA sequences or haplotypes produced by RFLP are short. It is based on the 1989 Tajima test, using the model of infinite sites without recombination. It commutes two estimators using the theta mutation as a parameter.

#### FS FU Test of Selective Neutrality

Also based on the model of infinite sites without recombination, the FU test is suitable for short DNA sequences or haplotypes produced by RFLP. However, in this case, it assesses the observed probability of a randomly neutral sample with a number of alleles equal to or less than the observed value. In this case the estimator used is θ.

### FOR METHODS THAT MEASURE INTERPOPULATION DIVERSITY

#### Genetic structure of the population inferred by molecular variance analysis (AMOVA)

This stage is the most used in the LaBECom protocols because it allows to know the genetic structure of populations measuring their variances, this methodology, first defined by Cockerham in 1969 and 1973) and, later adapted by other researchers, is essentially similar to other approaches based on analyses of gene frequency variance, but takes into account the number of mutations between haplotypes. When the population group is defined, we can define a particular genetic structure that will be tested, that is, we can create a hierarchical analysis of variance by dividing the total variance into covariance components by being able to measure intra-individual differences, interindividual differences and/or interpopulation allocated differences.

#### Minimum Spanning Network (MSN) among haplotypes

In LaBECom, this tree is generated using the operational taxonomic units (OTU). This tree is calculated from the matrix of paired distances using a modification of the algorithm described in Rohlf (1973).

#### Locus-by-locus AMOVA

We performed this analysis for each locus separately as it is performed at the haplotypic level and the variance components and f statistics are estimated for each locus separately generating in a more global panorama.

#### Paired genetic distances between populations

This is the most present analysis in the work of LaBECom. These generate paired F_ST_ parameters that are always used, extremely reliably, to estimate the short-term genetic distances between the populations studied, in this model a slight algorithmic adaptation is applied to linearize the genetic distance with the time of population divergence (Reynolds et al. 1983; Slatkin, 1995).

#### Reynolds Distance (Reynolds et al. 1983)

Here we measured how much pairs of fixed N-size haplotypes diverged over t generations, based on F_ST_ indices.

#### Slatkin’s linearized F_ST’s_ (Slatkin 1995)

We used this test in LaBECom when we want to know how much two Haploid populations of N size diverged t generations behind a population of identical size and managed to remain isolated and without migration. This is a demographic model and applies very well to the phylogeography work of our Laboratory.

#### Nei’s average number of differences between populations

In this test we assumed that the relationship between the gross (D) and liquid (AD) number of Nei differences between populations is the increase in genetic distance between populations (Nei and Li, 1979).

#### Relative population sizes: divergence between populations of unequal sizes

We used this method in LaBECom when we want to estimate the time of divergence between populations of equal sizes (Gaggiotti and Excoffier, 2000), assuming that two populations diverged from an ancestral population of N0 size a few t generations in the past, and that they have remained isolated from each other ever since. In this method we assume that even though the sizes of the two child populations are different, the sum of them will always correspond to the size of the ancestral population. The procedure is based on the comparison of intra and inter populational (π’s) diversities that have a large variance, which means that for short divergence times, the average diversity found within the population may be higher than that observed among populations. These calculations should therefore be made if the assumptions of a pure fission model are met and if the divergence time is relatively old. The results of this simulation show that this procedure leads to better results than other methods that do not take into account unequal population sizes, especially when the relative sizes of the daughter populations are in fact unequal.

#### Accurate population differentiation tests

We at LaBECom understand that this test is an analog of fisher’s exact test in a 2×2 contingency table extended to a rxk contingency table. It has been described in Raymond and Rousset (1995) and tests the hypothesis of a random distribution of k different haplotypes or genotypes among r populations.

#### Assignment of individual genotypes to populations

Inspired by what had been described in Paetkau et al (1995, 1997) and Waser and Strobeck (1998) this method determines the origin of specific individuals, knowing a list of potential source populations and uses the allelic frequencies estimated in each sample from their original constitution.

#### Detection of loci under selection from F-statistics

We use this test when we suspect that natural selection affects genetic diversity among populations. This method was adapted by Cavalli-Sforza in 1996 from a 1973 work by Lewontin and Krakauer.

## 2. Results

### Molecular Variance Analysis (AMOVA) and Genetic Distance

Genetic distance and molecular variation (AMOVA) analyses were not significant for the groups studied, presenting a variation component of 0.12 between populations and 4.46 within populations. The F_ST_ value (0.03) showed a low fixation index, with non-significant evolutionary divergences within and between groups, With a representative exception for haplotypes from Peru and Uruguai (Table 1) (Figures 1 and 2).

**Table 1.** Components of haplotypic variation and paired F_ST_ value for the 38 complete genome sequences of SARS-CoV-2 from South America.

| Source of variation | d.f. | Sum of squares | Variance components | Percentage of variation |
| --- | --- | --- | --- | --- |
| Among populations | 5 | 25.399 | 0.11704 $V_a$ | 2.56 |
| Within populations | 32 | 142.601 | 4.45630 $V_b$ | 97.44 |
| Total | 37 | 168.000 | 4.57334 |  |
| Fixation Index | **$F_{ST}$:** | **0.02559** |  |  |
| Significance tests (1023 permutations) |  |  |  |  |
| Va and FST: P (rand. value > obs. value) = 0.30010 |  |  |  |  |
| P (rand. value = obs. value) = 0.00000 |  |  |  |  |
| P-value = 0.30010+-0.01283 |  |  |  |  |

**Figure 1.**
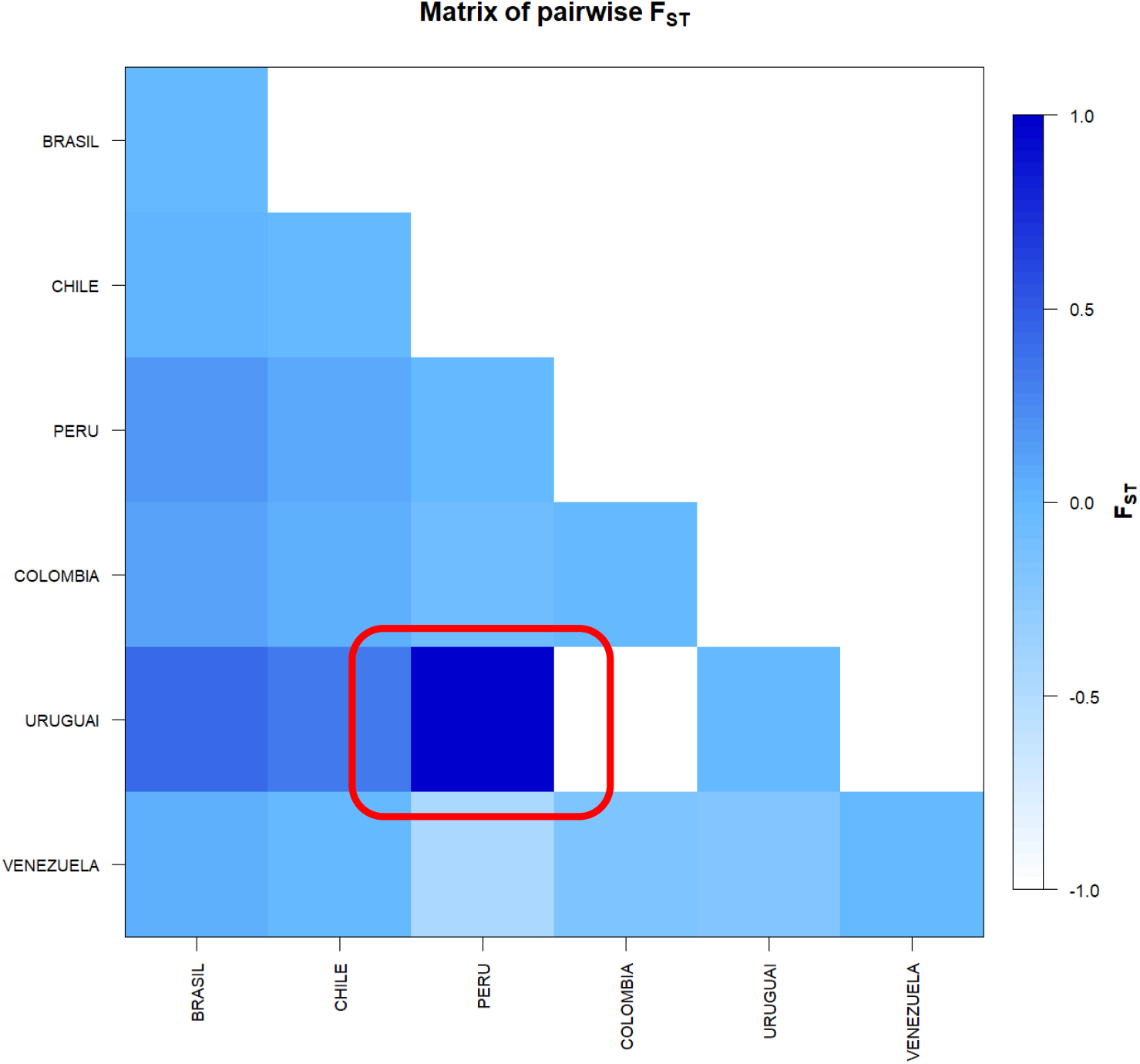
F_ST_-based genetic distance matrix between for the complete genome sequences of SARS-CoV-2 from six countries in South America. * Generated by the statistical package in R language using the output data of the Software Arlequin version 3.5.1.2

**Figure 2.**
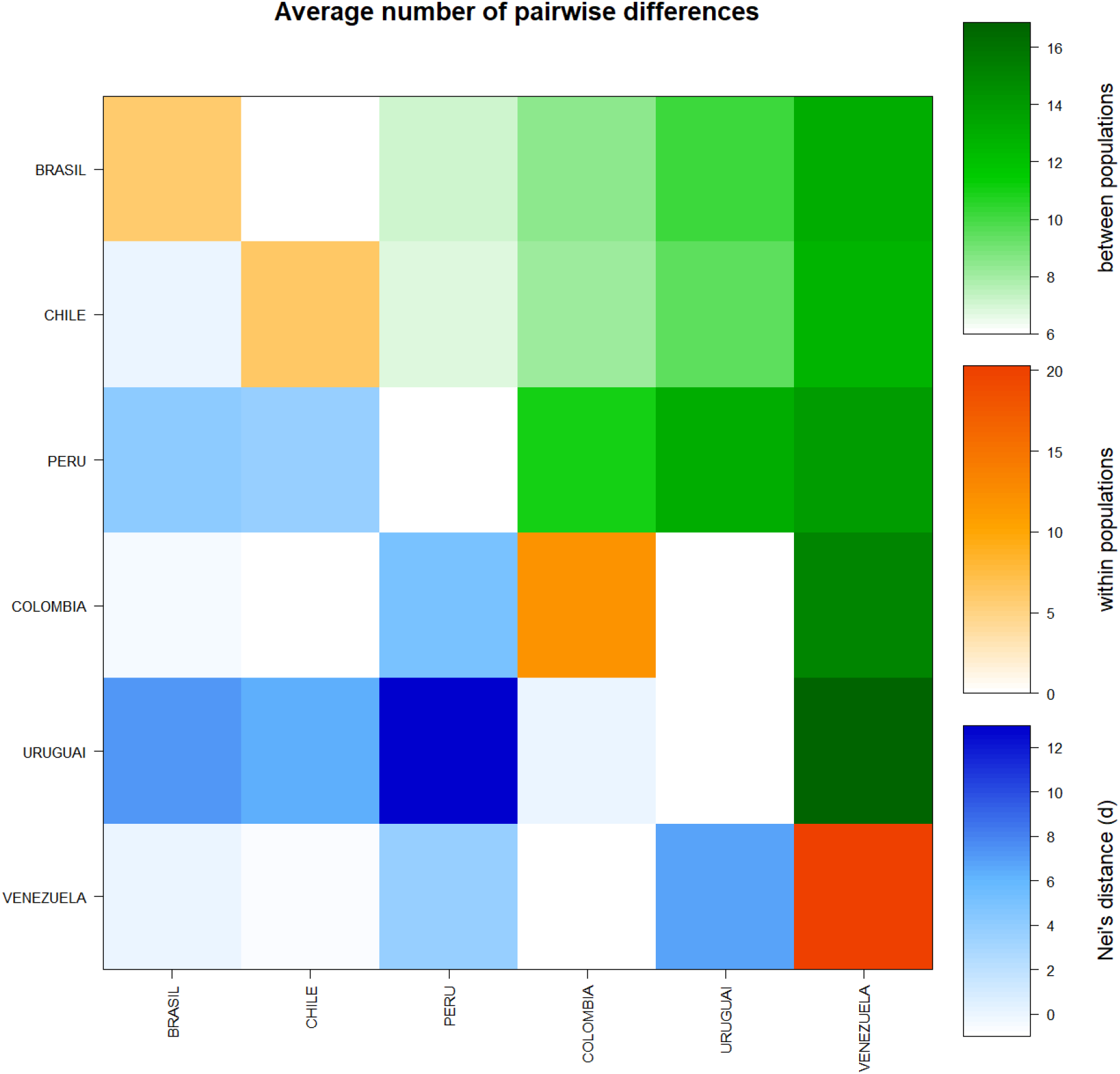
Matrix of paired differences between the populations studied: between the groups; within the groups; and Nei distance for the complete genome sequences of SARS-CoV-2 from six countries in South America.

A significant similarity was also evidenced for the time of genetic evolutionary divergence among all populations; supported by τ variations, mismatch analyses and demographic and spatial expansion analyses. With a representative exception for haplotypes from Venezuela (Table 2), (Figures 3, 4 5 and 6).

**Table 2.** Demographic and spatial expansion simulations based on the τ, θ, and M indices of sequences of the complete SARS-CoV-2 genomes from six South American countries.

| Statistics | BRASIL | CHILE | PERU | COLOMBIA | URUGUAI | VENEZUELA | Mean | s.d. |
| --- | --- | --- | --- | --- | --- | --- | --- | --- |
| **Demographic expansion** |  |  |  |  |  |  |  |  |
| Tau | 8.65821 | 3.41406 | 0.00000 | 0.00000 | 0.00000 | 8.00000 | 3.34538 | 4.08585 |
| Tau qt 2.5% | 1.43937 | 0.00000 | 0.00000 | 0.00000 | 0.00000 | 4.43744 | 0.97947 | 1.78922 |
| Tau qt 5% | 2.76561 | 2.33788 | 0.00000 | 0.00000 | 0.00000 | 5.36523 | 1.74479 | 2.17413 |
| Tau qt 95% | 12.31057 | 17.19734 | 0.00000 | 0.00000 | 0.00000 | 20.92787 | 8.40596 | 9.60534 |
| Tau qt 97.5% | 13.72265 | 18.72268 | 0.00000 | 0.00000 | 0.00000 | 21.95513 | 9.06674 | 10.27271 |
| Theta0 | 0.00000 | 4.28554 | 0.00000 | 0.00000 | 0.00000 | 5.49999 | 1.63092 | 2.55563 |
| Theta0 qt 2.5% | 0.00000 | 0.00000 | 0.00000 | 0.00000 | 0.00000 | 0.00000 | 0.00000 | 0.00000 |
| Theta0 qt 5% | 0.00000 | 0.00000 | 0.00000 | 0.00000 | 0.00000 | 0.00000 | 0.00000 | 0.00000 |
| Theta0 qt 95% | 1.82648 | 3.26617 | 0.00000 | 0.00000 | 0.00000 | 12.13044 | 2.87052 | 4.72678 |
| Theta0 qt 97.5% | 2.83189 | 4.79179 | 0.00000 | 0.00000 | 0.00000 | 16.98018 | 4.10064 | 6.60932 |
| Theta1 | 8.67921 | 13.59864 | 0.00000 | 0.00000 | 0.00000 | 3414.97837 | 572.87604 | 1392.35166 |
| Theta1 qt 2.5% | 3.11981 | 3.91268 | 0.00000 | 0.00000 | 0.00000 | 27.98066 | 5.83553 | 10.98763 |
| Theta1 qt 5% | 4.26270 | 4.56568 | 0.00000 | 0.00000 | 0.00000 | 39.51658 | 8.05749 | 15.56301 |
| Theta1 qt 95% | 33.99159 | 82.50502 | 0.00000 | 0.00000 | 0.00000 | 323.11280 | 73.26823 | 126.61357 |
| Theta1 qt 97.5% | 52.81974 | 162.02985 | 0.00000 | 0.00000 | 0.00000 | 590.61070 | 134.24338 | 232.26574 |
| SSD | 0.04330 | 0.00540 | 0.00000 | 0.00000 | 0.00000 | 0.07165 | 0.02006 | 0.03041 |
| Model (SSD) p-value | 0.30000 | 0.99000 | 0.00000 | 0.00000 | 0.00000 | 0.04000 | 0.22167 | 0.39418 |
| **Raggedness index** | **0.07035** | **0.00860** | **0.00000** | **0.00000** | **0.00000** | **0.16780** | **0.04112** | 0.06787 |
| Raggedness p-value | 0.38000 | 1.00000 | 0.00000 | 0.00000 | 0.00000 | 0.22000 | 0.26667 | 0.39144 |
| **Spatial expansion** |  |  |  |  |  |  |  |  |
| Tau | 6.18844 | 2.25056 | 0.00000 | 0.00000 | 0.00000 | 8.24067 | 2.77994 | 3.60283 |
| Tau qt 2.5% | 1.28581 | 0.69166 | 0.00000 | 0.00000 | 0.00000 | 3.44275 | 0.90337 | 1.34817 |
| Tau qt 5% | 3.98499 | 1.97354 | 0.00000 | 0.00000 | 0.00000 | 4.37483 | 1.72223 | 2.05513 |
| Tau qt 95% | 10.32285 | 13.44850 | 0.00000 | 0.00000 | 0.00000 | 14.53488 | 6.38437 | 7.12916 |
| Tau qt 97.5% | 10.82249 | 17.56023 | 0.00000 | 0.00000 | 0.00000 | 15.32114 | 7.28398 | 8.26907 |
| Theta | 1.64652 | 4.96606 | 0.00000 | 0.00000 | 0.00000 | 5.15534 | 1.96132 | 2.48474 |
| Theta qt 2.5% | 0.00072 | 0.00072 | 0.00000 | 0.00000 | 0.00000 | 0.00072 | 0.00036 | 0.00040 |
| Theta qt 5% | 0.00072 | 0.00072 | 0.00000 | 0.00000 | 0.00000 | 0.00258 | 0.00067 | 0.00100 |
| Theta qt 95% | 2.75007 | 7.34736 | 0.00000 | 0.00000 | 0.00000 | 16.21983 | 4.38621 | 6.46833 |
| Theta qt 97.5% | 3.02024 | 7.64974 | 0.00000 | 0.00000 | 0.00000 | 20.05757 | 5.12126 | 7.90674 |
| M | 2.30868 | 11.22560 | 0.00000 | 0.00000 | 0.00000 | 8827.30237 | 1473.47278 | 3602.62866 |
| M qt 2.5% | 0.52435 | 0.86722 | 0.00000 | 0.00000 | 0.00000 | 20.59232 | 3.66398 | 8.30087 |
| M qt 5% | 0.82693 | 1.18551 | 0.00000 | 0.00000 | 0.00000 | 33.82173 | 5.97236 | 13.65272 |
| M qt 95% | 12.86742 | 110.88566 | 0.00000 | 0.00000 | 0.00000 | 2097.48882 | 370.20698 | 847.30175 |
| M qt 97.5% | 15.34689 | 191.55404 | 0.00000 | 0.00000 | 0.00000 | 5327.87210 | 922.46217 | 2159.51526 |
| SSD | 0.02288 | 0.00560 | 0.00000 | 0.00000 | 0.00000 | 0.07137 | 0.01664 | 0.02824 |
| Model (SSD) p-value | 0.77000 | 0.98000 | 0.00000 | 0.00000 | 0.00000 | 0.10000 | 0.30833 | 0.44562 |
| **Raggedness index** | **0.07035** | **0.00860** | **0.00000** | **0.00000** | **0.00000** | **0.16780** | **0.04112** | 0.06787 |
| Raggedness p-value | 0.68000 | 1.00000 | 0.00000 | 0.00000 | 0.00000 | 0.22000 | 0.31667 | 0.42641 |

**Figure 3.**
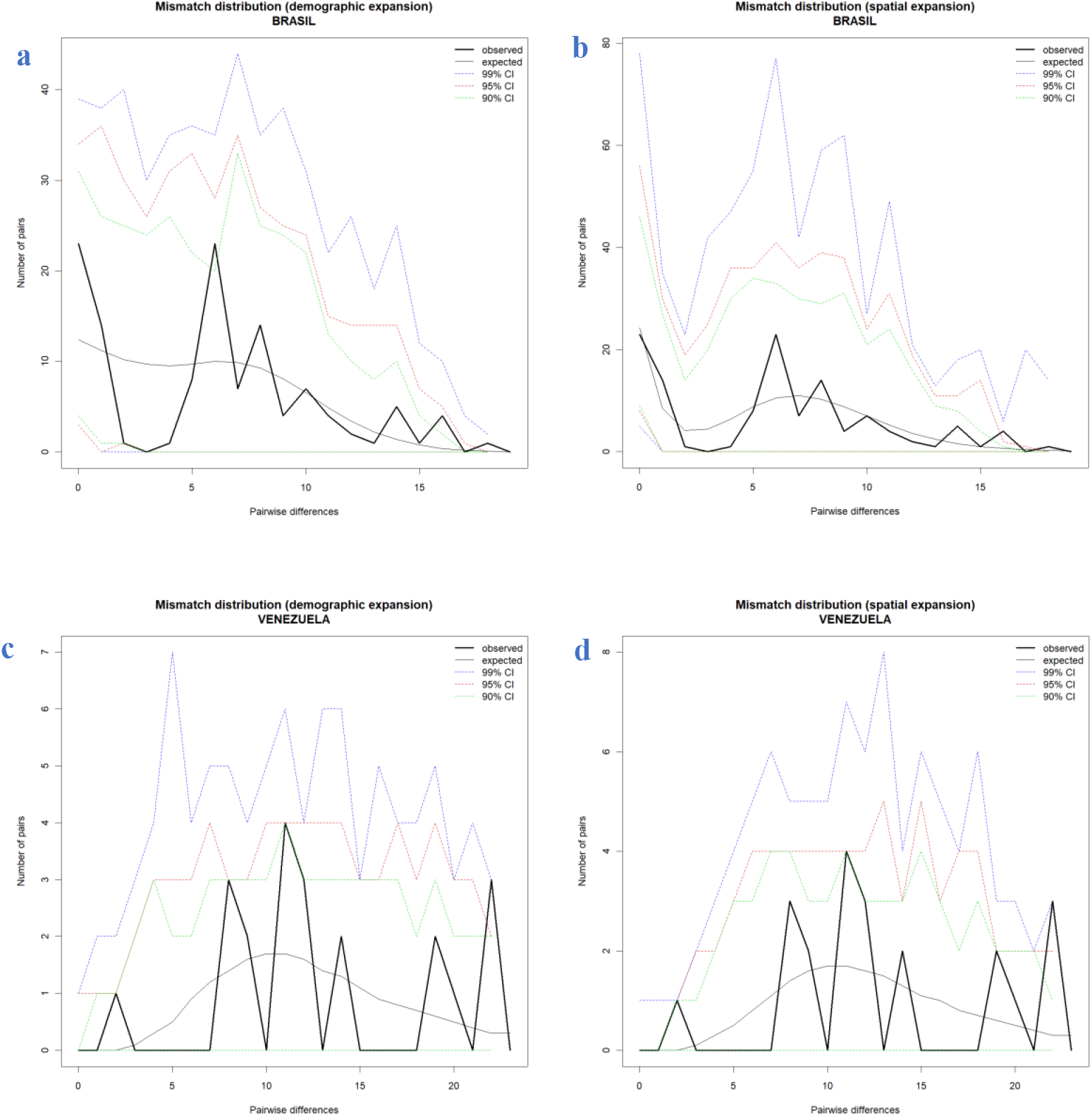
Comparison between the Demographic and Spatial Expansion of sequences of the complete genomes of SARS-CoV-2 from six countries in South America. (**a** and **b**) Graphs of demographic expansion and spatial expansion of haplotypes from Brazil, respectively; (**c** and **d**) Graphs of demographic expansion and spatial expansion of haplotypes from Venezuela, respectively. *Graphs Generated by the statistical package in R language using the output data of the Software Arlequin version 3.5.1.2

**Figure 4.**
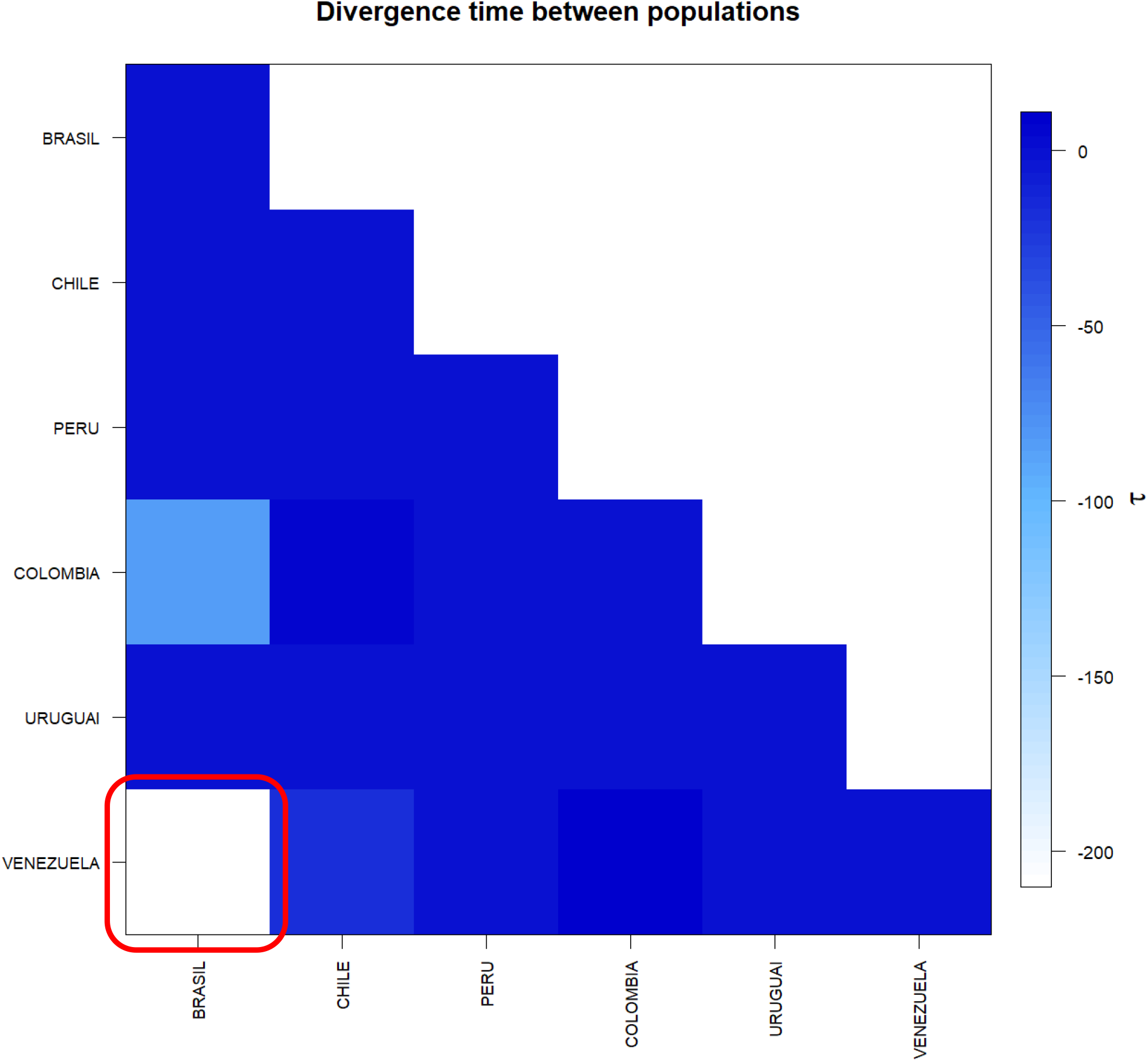
Matrix of divergence time between the complete genomes of SARS-CoV-2 from six countries in South America. In evidence the high value τ present between the sequences of Brazil and Venezuela. * Generated by the statistical package in R language using the output data of the Software Arlequin version 3.5.1.2.

**Figure 5.**
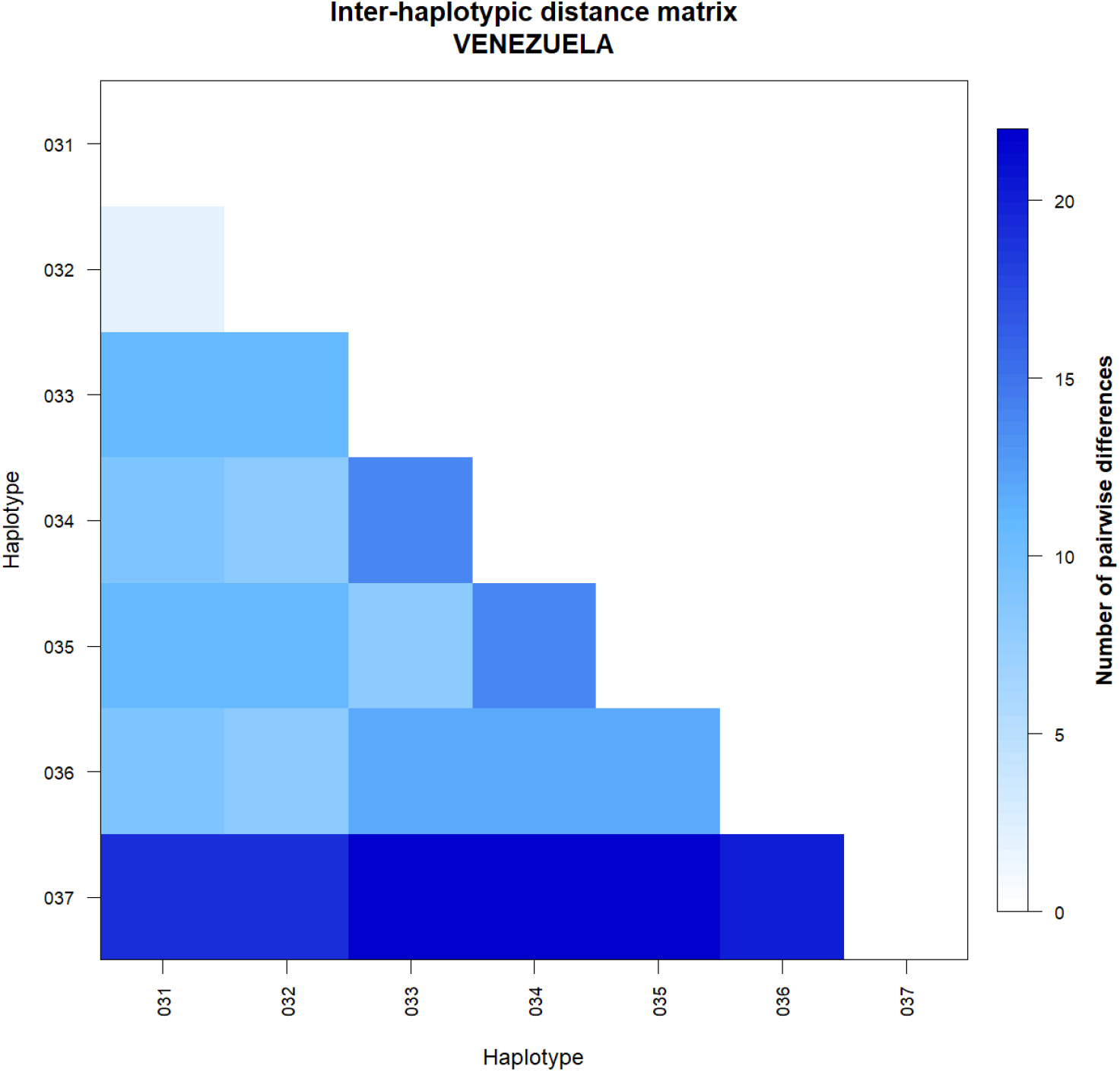
Matrix of inter haplotypic distance in the complete genomes of SARS-CoV-2 from Venezuela. **Note the great variation between haplotypes**. *Generated by the statistical package in R language using the output data of the Software Arlequin version 3.5.1.2.

**Figure 6.**
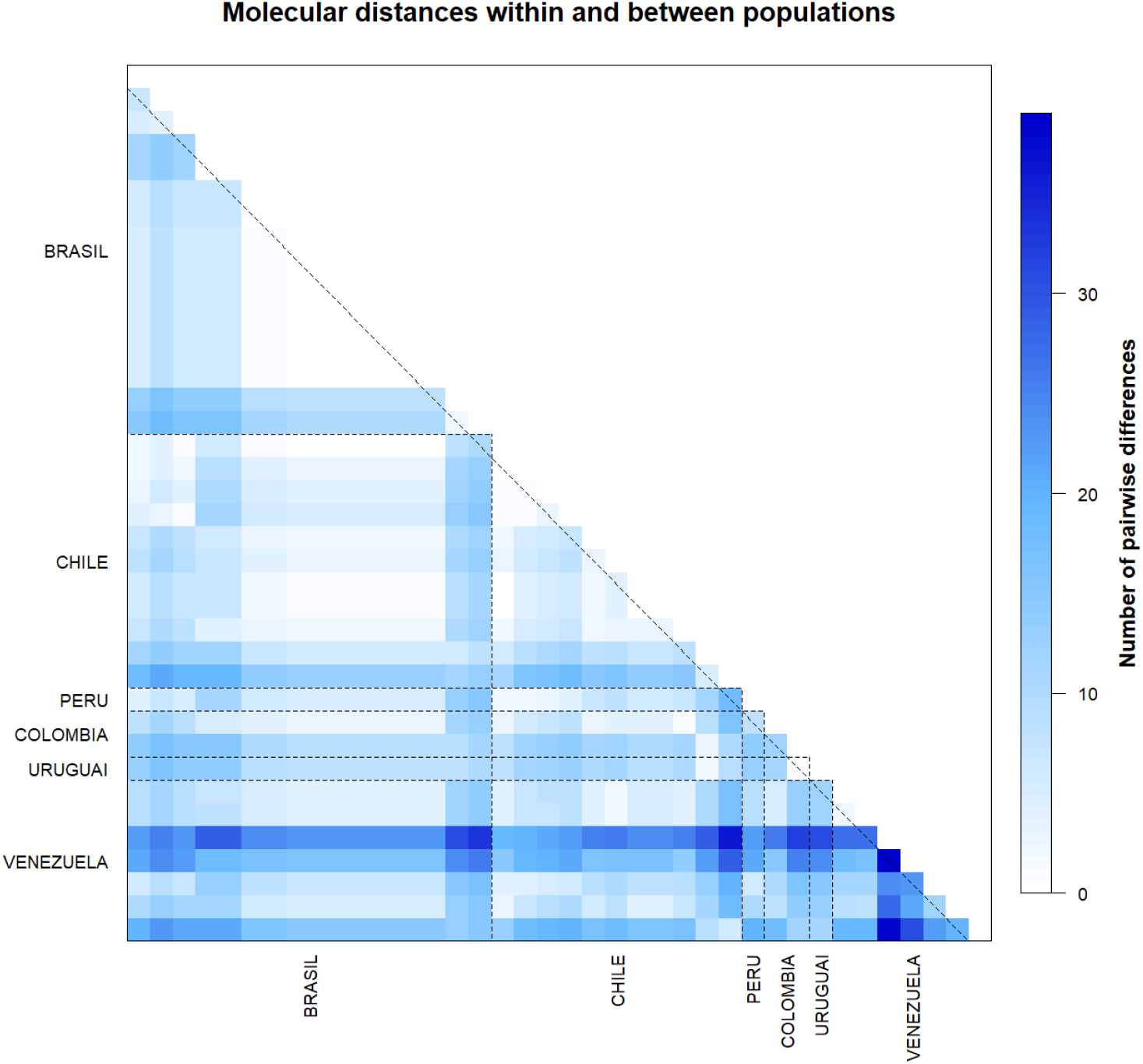
Matrix of inter haplotypic distance and number of polymorphic sites the complete genomes of SARS-CoV-2 from six countries in South America. **Note the great variation between haplotypes from Venezuela in relation to the others**. *Generated by the statistical package in R language using the output data of the Software Arlequin version 3.5.1.2.

The molecular diversity analyses estimated per θ reflected a significant level of mutations among all haplotypes (transitions and transversions). Indel mutations (insertions or additions) were not found in any of the six groups studied (Table 3). The D tests of Tajima and Fs de Fu showed disagreements between the estimates of general θ and π, but with negative and highly significant values, indicating, once again, an absence of population expansions (Table 4). The irregularity index (R= Raggedness) with parametric bootstrap, simulated new θ values for before and after a supposed demographic expansion and in this case assumed a value equal to zero for all groups (Table 2); (Figure 7).

**Table 3.**
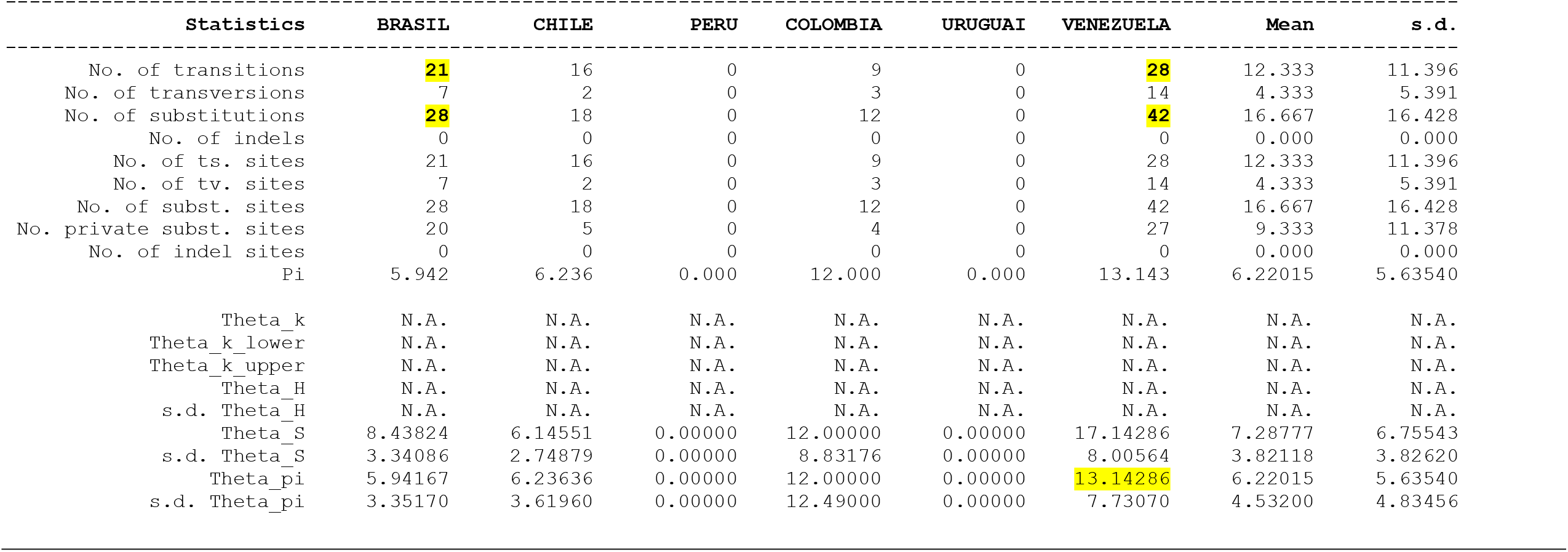
Molecular Diversity Indices for the complete Genomes of SARS-CoV-2 from six countries in South America

**Table 4.** Neutrality Tests for the complete Genomes of SARS-CoV-2 from six countries in South America

|  | Statistics | BRASIL | CHILE | PERU | COLOMBIA | URUGUAI | VENEZUELA | Mean | s.d. |
| --- | --- | --- | --- | --- | --- | --- | --- | --- | --- |
| Ewens-Watterson test |  |  |  |  |  |  |  |  |  |
|  | Sample size | 16 | 11 | 1 | 2 | 1 | 7 | 6.33333 | 6.18601 |
|  | No. of alleles(unchecked) | 16 | 11 | 1 | 2 | 1 | 7 | 6.33333 | 6.18601 |
|  | Observed F value | N.A. | N.A. | N.A. | N.A. | N.A. | N.A. | N.A. | N.A. |
|  | Expected F value | N.A. | N.A. | N.A. | N.A. | N.A. | N.A. | N.A. | N.A. |
|  | Watterson test: Pr(rand F <= obs F) | N.A. | N.A. | N.A. | N.A. | N.A. | N.A. | N.A. | N.A. |
|  | Slatkin's exact test P-value | N.A. | N.A. | N.A. | N.A. | N.A. | N.A. | N.A. | N.A. |
| Chakraborty's test |  |  |  |  |  |  |  |  |  |
|  | Sample size | 16 | 11 | 1 | 2 | 1 | 7 | 6.33333 | 6.18601 |
|  | No. of alleles(unchecked) | 16 | 11 | 1 | 2 | 1 | 7 | 6.33333 | 6.18601 |
|  | Obs. homozygosity | 0.00000 | 0.00000 | 0.00000 | 0.00000 | 0.00000 | 0.00000 | 0.00000 | 0.00000 |
|  | Exp. no. of alleles | 8.13974 | 6.67071 | 0.00000 | 1.92308 | 0.00000 | 5.78902 | 3.75376 | 3.56148 |
|  | P(k or more alleles) | N.A. | N.A. | N.A. | N.A. | N.A. | N.A. | N.A. | N.A. |
| Tajima's D test |  |  |  |  |  |  |  |  |  |
|  | Sample size | 16 | 11 | 1 | 2 | 1 | 7 | 6.33333 | 6.18601 |
|  | S | 28 | 18 | 0 | 12 | 0 | 42 | 16.66667 | 16.42762 |
|  | Pi | 5.94167 | 6.23636 | 0.00000 | 12.00000 | 0.00000 | 13.14286 | 6.22015 | 5.63540 |
|  | Tajima's D | -1.21891 | 0.06649 | 0.00000 | 0.00000 | 0.00000 | -1.34385 | -0.41604 | 0.67194 |
|  | Tajima's D p-value | 0.10600 | 0.57300 | 1.00000 | 1.00000 | 1.00000 | 0.07200 | 0.62517 | 0.44716 |
| Fu's FS test |  |  |  |  |  |  |  |  |  |
|  | No. of alleles(unchecked) | 16 | 11 | 1 | 2 | 1 | 7 | 6.33333 | 6.18601 |
|  | Theta_pi | 5.94167 | 6.23636 | 0.00000 | 12.00000 | 0.00000 | 13.14286 | 6.22015 | 5.63540 |
|  | Exp. no. of alleles | 8.13974 | 6.67071 | 0.00000 | 1.92308 | 0.00000 | 5.78902 | 3.75376 | 3.56148 |
|  | FS | -12.00112 | -6.00361 | 0.00000 | 2.48491 | 0.00000 | -1.09653 | -2.76939 | 5.31846 |
|  | FS p-value | 0.00000 | 0.00200 | N.A. | 0.56600 | N.A. | 0.18400 | N.A. | N.A. |

**Figure 7.**
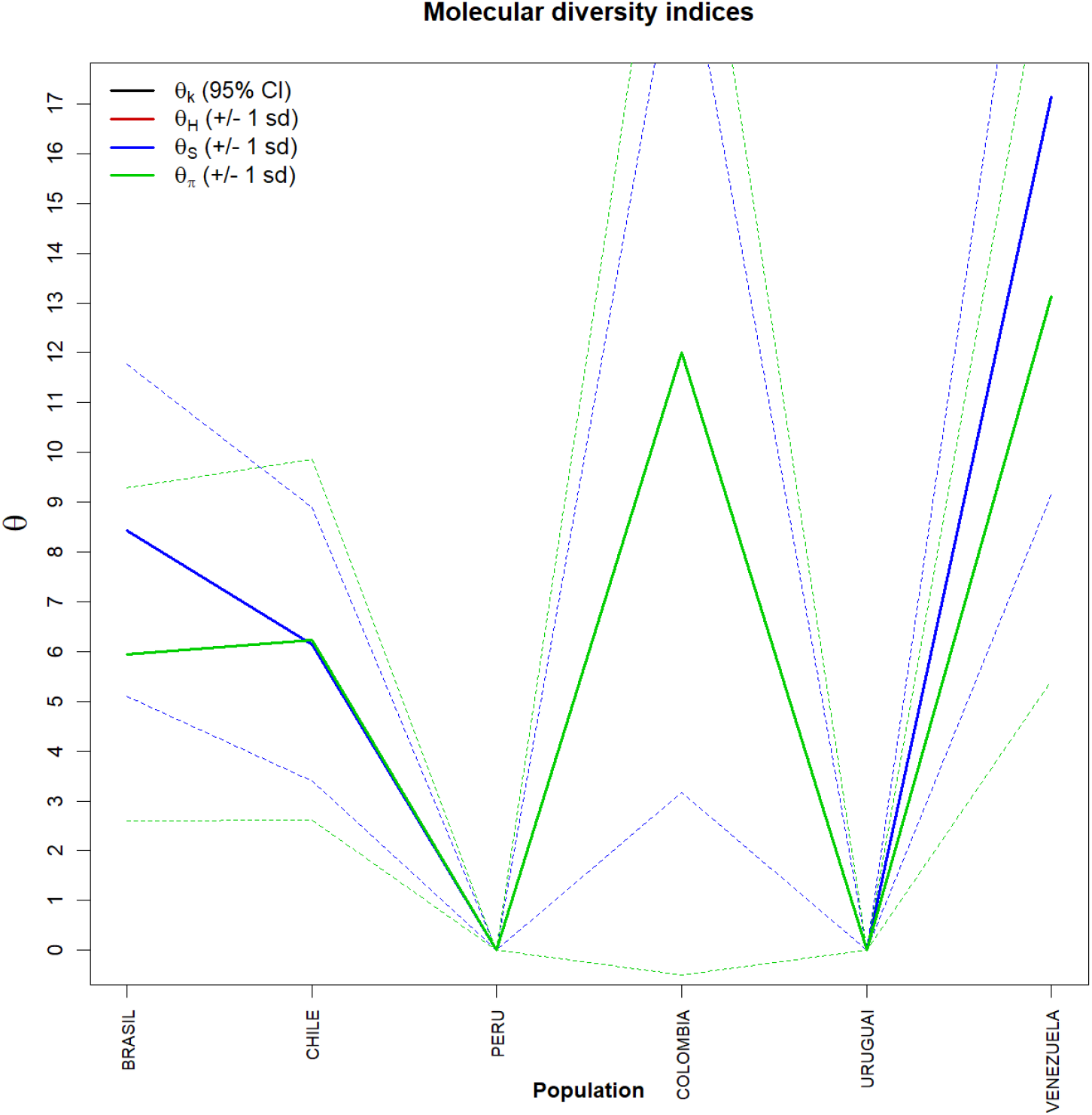
Graph of molecular diversity indices for the complete genomes of SARS-CoV-2 from six countries in South America. In the graph the values of θ: (θk) Relationship between the expected number of alllos (k) and the sample size; (θH) Expected homozygosity in a balanced relationship between drift and mutation; (θS) Relationship between the number of segregating sites (S), sample size (n) and non-recombinant sites; (θπ) Relationship between the average number of paired differences (π) and θ. * Generated by the statistical package in R language using the output data of the Arlequin software version 3.5.1.2.

## 5. Discussion

As the use of phylogenetic analysis and population structure methodologies had not yet been used in this PopSet, in this study it was possible to detect the existence of 6 distinct groups for the complete genome sequences of SARS-CoV-2 from South America, but with minimal variations among all of them. The groups described here presented minimum structuring patterns and were effectively slightly higher for the populations of Brazil and Venezuela. These data suggest that the relative degree of structuring present in these two countries may be related to gene flow. These structuring levels were also supported by simple phylogenetic pairing methodologies such as UPGMA, which in this case, with a discontinuous pattern of genetic divergence between the groups (supports the idea of possible sub-geographical isolations resulting from past fragmentation events), was observed a not so numerous amount of branches in the tree generated and with few mutational steps.

These few mutations have possibly not yet been fixed by drift by the lack of the founding effect, which accompanies the behavior of dispersion and/or loss of intermediate haplotypes throughout the generations. The values found for genetic distance support the presence of this continuous pattern of low divergence between the groups studied, since they considered important the minimum differences between the groups, when the haplotypes between them were exchanged, as well as the inference of values greater than or equal to that observed in the proportion of these permutations, including the p-value of the test.

The discrimination of the 38 genetic entities in their localities was also perceived by their small inter-haplotypic variations, hierarchised in all covariance components: by their intra- and inter-individual differences or by their intra- and intergroup differences, generating a dendogram that supports the idea that the significant differences found in countries such as Brazil and Venezuela, for example, were shared more in their form than in their number, since the result of estimates of the average evolutionary divergence found within these and other countries, even if they exist, were very low.

Based on the high level of haplotypic sharing, tests that measure the relationship between genetic distance and geographic distance, such as the Mantel test, were dispensed in this Estimators θ, even though they are extremely sensitive to any form of molecular variation (FU, 1997), supported the uniformity between the results found by all the methodologies employed, and can be interpreted as a phylogenetic confirmation that there is a consensus in the conservation of the SARS-CoV-2 genome in the Countries of America of America of South objects of this study, being therefore safe to affirm that the small number of existing polymorphisms should be reflected even in all their protein products. This consideration provides the safety that, although there are differences in the haplotypes studied, these differences are minimal in geographically distinct regions and thus it seems safe to extrapolate the levels of polymorphism and molecular diversity found in the samples of this study to other genomes of other South American countries, reducing speculation about the existence of rapid and silent mutations that, although they exist as we have shown in this work, they can significantly increase the genetic variability of the Virus, making it difficult to work with molecular targets for vaccines and drugs in general.

## Data Availability

The data that support the findings of this study are available from the corresponding author, Felix, P.T, upon request.

https://www.ncbi.nlm.nih.gov/labs/virus/vssi/#/virus?SeqType_s=Nucleotide&VirusLineage_ss=SARS-CoV-2,%20taxid:2697049&Completeness_s=complete&Region_s=South%20America

## Notes

### Competing Interest Statement

The authors have declared no competing interest.

### Funding Statement

This research did not receive any specific grant from funding agencies in the public, commercial, or not-for profit sectors.

### Author Declarations

This study does not contain any studies with human participants or animals performed by any of the authors.

